# “I didn’t come into nursing to be attacked and constantly abused”: A qualitative study of exposure to violence for nurses across ethnic groups in the United Kingdom

**DOI:** 10.1101/2024.05.02.24306749

**Authors:** Zoe Chui, Emma Caton, Habib Naqvi, Edward Baker, Juliana Onwumere, Geraldine A Lee, Stephani L Hatch

**Author notes:** Zoe Chui, E3.18, Institute of Psychiatry, Psychology & Neuroscience, 16 De Crespigny Park, London, UK SE5 8AB.

## Abstract

**Background:** Workplace violence is a serious threat to staff safety and leads to mental and physical health problems that have negative consequences for the recruitment and retention of nurses, amid the worst staffing crisis in the history of the National Health Service (NHS) in the United Kingdom.

**Objectives:** This study aims to explore the social context of violence for hospital-based and community nurses from different ethnic groups, the types of violence experienced or witnessed both in and outside the workplace, and the impact of violence on mental and physical health.

**Methods:** Semi-structured interviews were conducted online with 12 hospital-based and community nurses recruited across London. Interview data were analysed using reflexive thematic analysis.

**Results:** The sample comprised seven hospital nurses and five community nurses. Four themes were identified from the analysis: i) the social context in which nurses from different ethnic groups are exposed to community violence; ii) the types of workplace violence experienced or witnessed by hospital-based and community nurses from different ethnic groups; iii) nurses’ perceptions of the factors contributing to workplace violence iv) how violence impacts mental and physical health outcomes for hospital-based and community nurses from different ethnic groups. Based on the social ecological framework and the sociological theory of stress, we have used these findings to present a conceptual stress process model of violence exposure for nurses.

**Conclusions:** Hospital-based and community nurses from different ethnic groups are exposed to violence both in and outside the workplace which negatively affects their mental and physical health. Whilst interventions aimed at improving personal safety and security measures in hospitals are valuable, interventions to address the social and institutional factors that put nurses at risk of violence exposure are needed. Further research using wider criteria for violence to include witnessing and hearing about violent events are needed to advance our understanding of how nurses are affected by multiple sources and types of violence in both their work and personal lives.

**What is already known:**

- Workplace violence is widespread and can lead to mental and physical health problems for healthcare staff.
- Nurses are one of the occupational groups most vulnerable to workplace violence and are consistently in short supply.

**What this paper adds:**

- Findings informed the development of a conceptual stress process model of violence exposure for nurses.
- Nurses perceived their gender, age and ethnicity as contributing factors to workplace violence.
- Witnessing or experiencing violence both in and outside the workplace has serious negative consequences at the individual and organisational levels.

## Background

Workplace violence in healthcare is widespread and affects staff recruitment and retention, and patient care [1-3]. The World Health Organisation defines violence as “the intentional use of physical force or power, threatened or actual, against oneself, another person, or against a group or community, that either results in or has a high likelihood of resulting in injury, death, psychological harm, maldevelopment, or deprivation” [4]. Nurses are one of the occupational groups that are most at risk of workplace violence [5, 6] and are also consistently in short supply with 47,000 vacancies in the National Health Service (NHS) currently [7]. In 2021, the annual NHS Staff Survey reported that 14.3% of NHS staff in England experienced physical violence from patients, relatives or members of the public [8]. In light of the COVID-19 pandemic and its disproportionate impact on ethnic minority groups and frontline NHS staff, this study explores the impact of workplace violence on the mental and physical health of nurses in London, the majority of whom (56%) are from ethnic minority backgrounds [9].

Nurses experience many different types of violence including physical violence, verbal abuse, sexual harassment and threats [10-13]. Patients and their family members tend to be the main identified perpetrators but there are instances of staff-on-staff violence in some cases [14]. Exposure to workplace violence may be direct through first-hand experience, indirect through second-hand witnessing, or both [15].

Workplace violence is associated with mental health (e.g., depression and anxiety) [16, 17] and physical health problems (e.g., back pain, somatic symptoms and musculoskeletal disorder symptoms) [18]. Other consequences include workplace absenteeism and sickness, poor relationships with colleagues and avoidance of the workplace [19-21], which are detrimental to recruitment and retention. This is particularly damaging for specialised areas such as emergency care, critical care and mental health, which have a chronic shortage of experienced nurses.

While the mental and physical effects of direct experiences of violence are well-established [22, 23], less is known about the impact of witnessing violence and whether cumulative effects exist for those who experience both types of exposure. Furthermore, the majority of nursing research in the UK has focused on workplace violence exposure for nurses working in mental health wards or accident and emergency departments. However, more attention needs to be paid to community nurses who work in the intimate setting of a person’s home and regularly enter unfamiliar neighbourhoods.

With crime rates rising across London [24], community nurses may be further exposed to violence in their own communities. Studies of ethnic minority women living in urban areas have found that witnessing community violence is associated with poor mental health outcomes, and those directly and indirectly exposed to multiple sources of violence have poorer outcomes [25, 26]. It is therefore important to explore the cumulative impact of violence experienced or witnessed, both in the workplace and in the community, on mental and physical health outcomes for nurses from different ethnic groups.

Using semi-structured qualitative interviews with hospital-based and community nurses from different ethnic groups, this study aims to explore the social context of violence, the types of violence experienced or witnessed both in and outside the workplace, and the impact of violence on mental and physical health.

## Theoretical framework

The social context of violence exposure for healthcare workers is complex and can be understood using the social ecological framework recommended by the World Health Organisation Violence Prevention Alliance, which views interpersonal violence as the outcome of interactions between multiple risk factors at four levels: societal, institutional, interpersonal and individual [27, 28]. Societal factors that facilitate or prevent violence include economic and social policies that perpetuate structural inequalities. In the UK, over a decade of austerity and vast public spending cuts to health and social care services have led to an overburdened health system and chronic staff shortages [29, 30]. Other societal factors include social and cultural norms that condone violence such as accepting workplace violence in healthcare as being an unfortunate but inevitable “part of the job” [31, 32].

Risk factors for violence exposure at the institutional level include insufficient support from organisational leaders and a lack of violence prevention policy, education and program evaluation [31]. At the interpersonal level, personal relationships and social interactions influence the risks of becoming a victim or perpetrator of violence and include poor communication and distrust between healthcare practitioners and patients, and bullying behaviours between colleagues [31]. Finally, at the individual level, risk factors for violence exposure include insufficient awareness of prevention measures, underreporting violent incidents to prevent future occurrences and patients’ sociodemographic characteristics, expectations and satisfaction [31].

Exposure to violence is stressful in and of itself and can lead directly to poor mental and physical health outcomes. However, according to Pearlin’s sociological study of stress, violence exposure can also indirectly impact mental and physical health outcomes through the process of stress proliferation whereby the violent incident itself, which is the primary stressor, can lead to secondary stressors such as chronic role strain [33]. Despite being classed as “secondary”, these stressors may have an equal or even greater effect on health outcomes than the primary stressors due to their repeated and enduring nature, and the importance and value that people assign to these roles [33].

Crucially, the extent to which people are affected by violence exposure differs greatly depending on their social and personal resources. Within the workplace, social resources may take the form of emotional and social support from co-workers or leaders in the organisation to adequately address and prevent workplace violence [34, 35]. Personal resources may include the extent to which one’s sense of control buffers the impact of job demands on mental strain and also increases job satisfaction by providing an intellectually stimulating environment to learn new skills and develop competency [36]. However, personal resources, and one’s ability to cope with the primary and secondary stressors, may be depleted by cumulative exposure to multiple violent incidents [37, 38].

## Methods

### Study design

This is a cross-sectional, qualitative study using semi-structured interviews. This study partners with NHS Race and Health Observatory (RHO) and builds on the Wellcome and ESRC funded Tackling Inequalities and Discrimination Experiences in health Services (TIDES) study which investigates how discrimination experienced by both patients and healthcare practitioners may generate and perpetuate inequalities in health services (www.tidesstudy.com).

### Participants

To be eligible for the study, participants needed to be in permanent employment within the National Health Service (NHS), in either hospital-based or community-based nursing roles, and have experienced or witnessed at least one violent incident (as defined by the World Health Organisation [4]) in the workplace since qualifying as nurses. Due to differences in employment and management policies, we excluded student nurses or nursing associates, those without a substantive contract in a London-based NHS Trust (e.g., retired, temporary “bank” or private sector nurses) and those without current Nursing and Midwifery (NMC) registration. For the purposes of our study objectives, we excluded those who had never experienced or witnessed violence in the workplace, or who experienced workplace violence in a different occupational position.

Participants were recruited between May and August 2021 via email circulars and social media posts sent by NHS England, the Royal College of Nursing (RCN), Freedom to Speak Up (FTSU) Guardians at individual NHS Trusts, and by re-contacting participants and collaborators from the TIDES study. Nurses interested in participating contacted the researcher directly and eligibility was assessed via email correspondence. To mitigate pressures to participate, participants were given at least 48 hours to read the information sheet before being re-contacted.

### Procedure

Prior to the interview, participants completed a consent form, sociodemographic characteristics form and contact details form using online survey tool Qualtrics (www.qualtrics.com). The semi-structured interviews were conducted by a graduate-level researcher via video conferencing software Microsoft Teams due to the ongoing pandemic. The interviews were approximately 60 minutes in duration, using a topic guide developed from the literature and with guidance from a nurse academic. The interviews began with a broad question about the participant’s current role and key elements of their job. Participants were then asked about violent incidents that they had personally experienced or witnessed in the workplace, and the impact, if any, that these incidents had on their emotional wellbeing and ability to carry out their work. Finally, to explore the cumulative effect of violence exposure, participants were asked about violent incidents they had personally experienced or witnessed outside of the workplace and how safe they felt in the area they lived.

Once the interview was completed, participants were compensated for their time with a £15 voucher, signposted to support services and, where appropriate, offered one-on-one debrief sessions with the study clinician. Each interview was digitally recorded to ensure accuracy and then transcribed verbatim by external transcription companies that had signed data handling and confidentiality agreements.

### Data analysis

Interview transcripts were assessed for accuracy then transferred to NVivo 12 [39] in order to apply codes to the data, sort and organise the data and facilitate data retrieval. Data were examined using the six stages of reflexive thematic analysis as outlined by Braun and Clarke [40]: 1) the text was read multiple times to become familiar with the content and get an understanding of what participants said and issues that arose; 2) the entire dataset was then coded with succinct labels to identify important information which might be relevant to the research question; 3) codes were then grouped together into broader themes; 4) themes were then checked against the original data set to ensure that they accurately represented the data; 5) themes were examined in detail to determine their meaning and generate an informative name; 6) finally, the analytic narrative was developed, extract examples were selected and the analysis was contextualised in relation to existing literature.

### Rigor and trustworthiness

Several strategies were used to ensure rigor and trustworthiness of our results. Two researchers (ZC and EC) independently codified key themes and then compared coding frameworks through a process of analysis triangulation. The researchers identified similar themes from the data which were discussed with the other authors. Professionals from different multidisciplinary backgrounds such as psychology, sociology, nursing, equality and diversity policy and public health (SH, GL, JO, EB, HN) advised on study design and commented on manuscript drafts. Participants did not provide feedback on the findings.

### Research team and reflexivity

ZC prepared study materials, recruited participants and conducted interviews. The researcher identifies as an East Asian woman who has a non-clinical background in health inequalities research and is external to the NHS. The other authors are from ethnically diverse backgrounds and hold a range of academic, clinical and/or public health roles. The research was conducted as part of ZC’s PhD at an academic institution which works in partnership with some London NHS Trusts. We were mindful of the impact this may have on participants’ concerns around confidentiality and recounting traumatic experiences to a non-clinical researcher. To mitigate this, the researcher explained the confidentiality protocol, signposted participants to support services and offered one-on-one debrief sessions with the study clinician after the interview. The researcher also kept a reflexive journal throughout data collection and analysis.

### Ethical approval

Ethical approval was granted by the King’s College London Research Ethics Committee for Psychiatry, Nursing and Midwifery (HR/DP-20/21-22048).

## Results

### Sample characteristics

The analytic sample comprised 12 nurses recruited from London-based NHS Trusts. All nurses who met the inclusion criteria, regardless of sociodemographic characteristics, were approached using the recruitment strategies outlined above. However, only female nurses volunteered to take part in the study. Over half of the participants (7/12) identified as belonging to a racial or ethnic minority group, almost all participants (11/12) identified as heterosexual and most participants (10/12) qualified after 2000. Seven participants worked in hospitals and five worked in community settings. Half the sample specialised in Mental Health Nursing, a third specialised in Child Nursing and the remainder specialised in either Adult or District/Community Nursing. Three-quarters of participants were highly senior nurses at Band 7 or 8 and the remainder were at entry level Band 5 or early senior roles at Band 6. One third of participants had worked for less than 12 months in their current position, half had worked for 1 to 5 years, and the remainder had worked in their current position for more than 11 years. Almost all participants (11/12) worked full-time.

### Key themes

Four key themes were conceptualized using Pearlin’s Stress Process Model [33] (see Figure 1). First, we described the social context in which nurses are exposed to violence both in the workplace and in the community. Second, we identified the types of violence experienced or witnessed (primary stressors) and how they lead to chronic role strains (secondary stressors). Finally, we examined the role of social support and cumulative adversity (social and personal resources) and described the subsequent mental and physical health outcomes.

**Figure 1.**
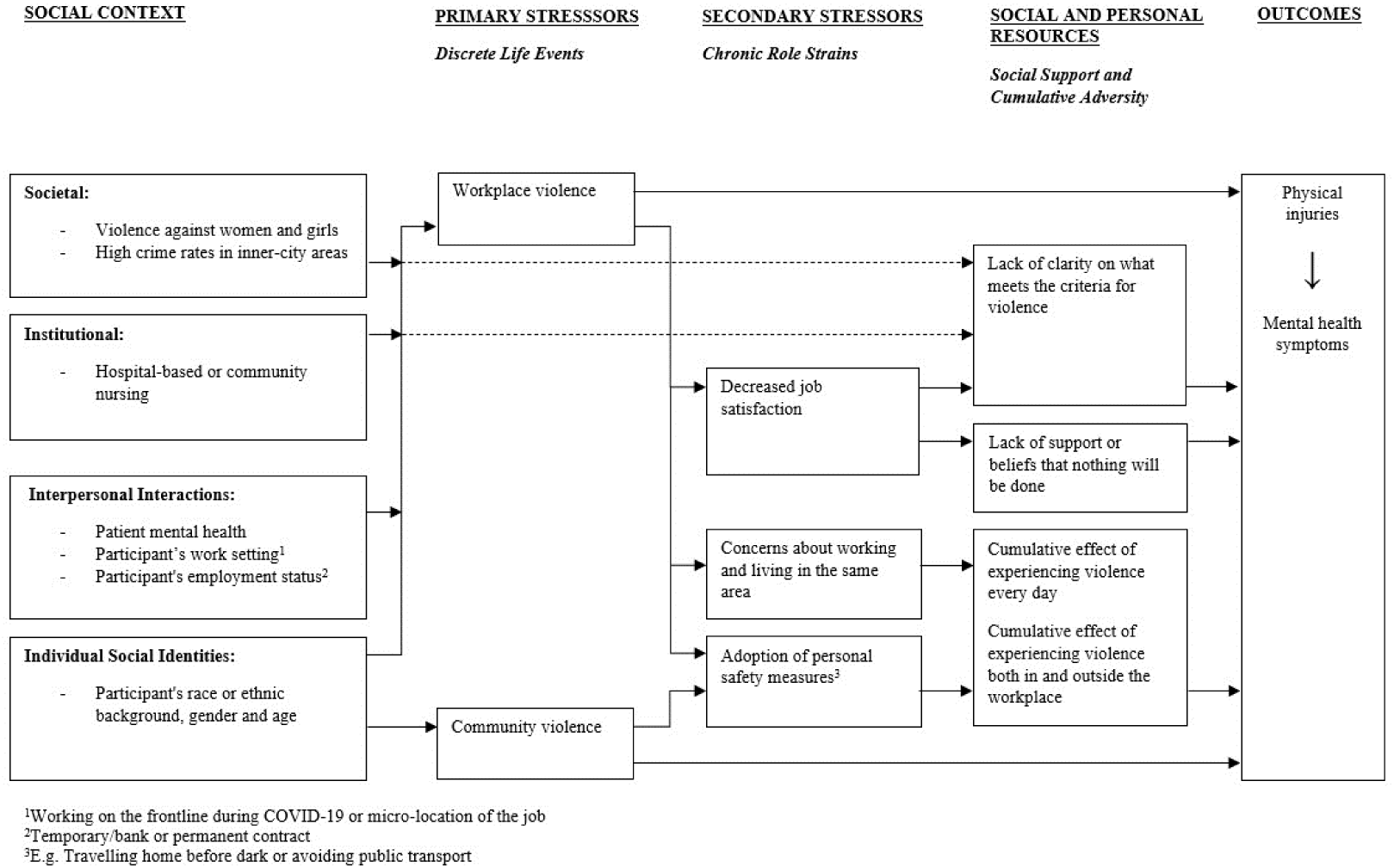
Conceptual stress process model of violence exposure for nurses

### The social context of violence against nurses

Participants reported experiences of violence at work in the context of their wider experiences of violence in their personal lives. The most common type of violence experienced outside of the workplace was harassment from men, especially when travelling alone at night.

> “Coming home in the dark on my own I would probably get a taxi […], more just from being a female and not wanting to be stopped by males.” Participant 8, Community Nurse, White British.

As well as direct experiences of violence, some participants reported traumatic experiences of witnessing violence towards their friends and members of their community. As a result, participants were more cautious about their own personal safety and more protective of their families.

> “I witnessed my best friend being stabbed at a party […]. He was stabbed twice in his chest and three times in his arm. I was involved as in I had to take his top off and wrap it around to stop the bleeding and stuff.” Participant 4, Community Nurse, White and Black Caribbean.

> “I did have a family friend who was stabbed and died a few years ago in South London. Which was awful. Protecting his daughter. So, I think that definitely had an impact on all the families and our children […]. It definitely made me a lot more cautious and had a bearing on my son and saying, ‘You need to be very careful with where you are going, and who you are mixing with.’” Participant 6, Hospital Nurse, White British.

At the institutional level, both hospital-based and community nurses were vulnerable to workplace violence. The frequency of violence was perceived by all participants to be higher in hospital settings due to patients in hospital being more acutely unwell than patients in the community. However, participants unanimously agreed that those in the community were more at risk of getting hurt by patients due to lone working and the lack of security measures typically found in hospitals.

> “I think the wards themselves you are exposed to more risk on a day-to-day basis but I think the risk of exposure in the community is higher because on a ward you’ve got someone there 24 hours a day and you’ve always got other staff around you to back you up, to support you. When you are in the community it’s just you and your care plan […]. So I think the risk is maybe a bit more extreme in the community because you are more lone working. You are going into patient’s houses on your own and not in pairs.” Participant 4, Community Nurse, White and Black Caribbean.

At the interpersonal level, workplace violence was attributed to patients’ mental health problems and consequently taken less seriously or justified by staff.

Furthermore, violence was seemingly tolerated more in mental health services compared to other services.

> “We are working with a group of really unwell people. Yes, violence should not be a part of your everyday, but people are unwell and that is what happens.” Participant 6, Hospital Nurse, White British.

> “It seems to feel sometimes that people with mental health [problems] can do a lot before anybody acts on it, whereas in other services I don’t think they would. I used to work in an addiction service and we had zero tolerance of abuse and threats, and the minute people did anything they were discharged. But in mental health services they don’t tend to do that so you have to continue to experience it until it gets quite excessive.” Participant 5, Hospital Nurse, White and Black Caribbean.

Another perceived factor contributing to workplace violence was the participant’s work setting. For example, nurses were seemingly more visible “on the frontline” compared to other healthcare practitioners so had a higher risk of experiencing violence. This was exacerbated by the COVID-19 pandemic and the additional challenges that patients and staff were required to manage, which meant that existing de-escalation techniques were less effective.

> “Because they’re [patients] coming with low resilience, they’re anxious, they’re stressed, everyone is fed up of the pandemic, it’s affected everyone differently I feel like they’re behaving a lot worse than they normally would.” Participant 3, Hospital Nurse, White British.

> “We’ve got the pandemic, people come in stressed, they’re scared of being out and about with people and we’ve had de-escalation training however I feel that’s not going to solve this. I know it can’t be solved but that’s not going to reduce it massively because a lot of people come at that pressure point […] and however nicely you put it, if you’re not saying yes to them, they’re going to explode.” Participant 3, Hospital Nurse, White British.

Employment status was also an important factor contributing to workplace violence because patients tried to “get away with a bit more” with temporary/bank staff compared to permanent staff.

> “I think our [bank] workers, our agency workers, are targeted slightly more because they are not a regular member of staff. A bit like how children are with a supply teacher in school. They think they can get away with a bit more and are less likely to have boundaries put in place because the worker will be less confident because they are not there all the time.” Participant 6, Hospital Nurse, White British.

At the individual level, the participants’ gender, age and ethnicity contributed to workplace violence for both hospital-based and community nurses.

> “So ethnicity and gender and age and sex all plays into it, yes. The patient that has the issue with me, he would never assault male staff, he only ever intimidates and threatens female staff generally. And definitely I think in some cultures, they still don’t respect women […]. Certain patients don’t want to be spoken to by an African person because there’s lots of racism from the patients […]. If a young nurse is trying to speak to an older patient they don’t always think that the younger nurse knows what they’re talking about.” Participant 5, Hospital Nurse, White and Black Caribbean.

## Types of violence

### Workplace violence (experienced)

Both hospital-based and community nurses experienced assault, aggression, threats or verbal abuse and/or sexual harassment. Perpetrators were either patients or patients’ relatives. Participants reported being assaulted by patients, including being smacked or punched, kicked, scratched, spat on, shoved, strangled, headbutted and hit with objects (e.g., mobile phones, cutlery or furniture).

> “Patients can lash out and become violent whilst they’re […] in bed and it is the nurses who are at risk of that. I have been kicked in the chest moving a patient between wards on a bed in the hospital before.” Participant 9, Hospital Nurse, White British.

Participants working in both settings reported aggression, threats and verbal abuse from both patients and patients’ relatives. This included shouting, swearing and racially abusing staff, following or chasing staff around the ward, filming staff and threatening to report them to the police or the media, threatening to wait outside for staff and beat them up, threatening to follow staff home or send someone to their home and threatening to kill staff.

> “Patients would often stand over me and threaten to kill me and threaten to want to come outside and beat me up.” Participant 5, Hospital Nurse, White and Black Caribbean.

Sexual harassment from patients included chasing and accosting staff and making sexually explicit comments and gestures.

> “One of the patients was quite a young male with a history of sexual assault. There was a particular occasion where he took down his trousers and his pants, and he was naked below the waist, and he started humping the wall and pointing at me, and then was telling me what he wanted to do to me, and how he was going to do it, and chased me down the corridor.” Participant 4, Community Nurse, White and Black Caribbean.

### Workplace violence (witnessed)

Witnessing violence was more commonly reported than experiencing violence in both hospital and community settings. Participants witnessed violence from patients towards staff and from patients towards other patients. Examples of violent incidents witnessed by participants include patients hitting, strangling and pinning staff to the ground. Participants also witnessed patients punching other patients or attacking them with metal objects.

> “Sometimes a patient will just come up and assault another patient. I think two weeks ago another patient just came off intensive care unit and went on the ward, within a few hours he had assaulted two patients […], two unprovoked attacks.” Participant 5, Hospital Nurse, White and Black Caribbean.

Accounts of patients harming themselves were more commonly reported by those working with young people than adults.

> “I think a lot more people end up hurting themselves, I guess. Which can seem quite violent as well, in various forms. So, on the ward I would have people coming up to me dripping with blood after they have cut themselves in various places.” Participant 8, Community Nurse, White British.

### Community violence

Direct experiences of violence outside the workplace were not common in this sample but some participants reported assault, aggression, sexual harassment, and racial abuse. Perpetrators were male strangers encountered in public spaces.

> “I have been a victim of assault. I was out drinking many years ago and somebody put something in my drink, and some guy dragged me across the street and was trying to steal my bag, and I don’t remember much of it but the guys chased him down from the pub, and he was saying he was going to kill me if they didn’t run away. That was quite traumatising actually.” Participant 5, Hospital Nurse, White and Black Caribbean.

Witnessing violence outside the workplace was less common than witnessing violence in the workplace in this sample. Some examples of violent incidents that were witnessed outside the workplace included fights between young people on public transport, knife crime and police violence.

> “I’d often see children getting on the bus attacking other kids or making threats to want to attack other kids and just somebody getting on a bus saying, I’m in a bad mood today I’m going to get someone, for no reason, and I couldn’t believe what I was hearing and I just thought, oh my God some poor child is going to get attacked today for doing nothing just because this person is unhappy.” Participant 5, Hospital Nurse, White and Black Caribbean.

> “Someone the other day was restrained by four policemen in my road, and that really worries me. I actually went out because they were kneeling on him and they had him face down […] and I was like, ‘You need to turn him over.’” Participant 6, Hospital Nurse, White British.

### Chronic role strains

Experiencing or witnessing violence in the workplace led to decreased job satisfaction and ultimately led some nurses to leave their roles and seek roles on a different ward/service.

> “I did have days that were really enjoyable, really rewarding and I’d go home feeling like I’d made a difference, but unfortunately all of that experience and trauma that’s associated with it shadowed a lot of the enjoyment that I could have gotten out of the job and that’s why I made the decision to walk away from that environment because I felt like if I didn’t it was either going to be my love for the job, my sanity or how I do the job and I’m not willing to compromise on any of that.” Participant 12, Hospital Nurse, Black African.

> “I didn’t come into nursing to be attacked, constantly abused.” Participant 5, Hospital Nurse, White and Black Caribbean.

Violence in the workplace also led to concerns about working and living in the same area, especially for those who had been threatened with violence by a patient or knew that the patient was part of a local gang, with implications for not reporting violence to the police.

> “I was constantly in fear of bumping into this patient, constantly wondering what if the people that she threatened to come after me even when I wasn’t at work would find their way to me, bearing in mind I don’t live too far from work.” Participant 12, Hospital Nurse, Black African.

> “Our local police officer came to take a statement and I did not feel comfortable reporting it because I was aware that she was involved with gangs in an area where I live, and I decided on balance that actually I just did not think it was worth it.” Participant 6, Hospital Nurse, White British.

Experiences of violence both within and outside of the workplace led to the adoption of personal safety measures such as taking a longer but better lit route home or avoiding public transport.

> “I have not been able to get onto public transport since October 2020, which I used to do quite easily, predominantly because of what is now an innate fear of either bumping into her or just I become even more hyper-vigilant.” Participant 12, Hospital Nurse, Black African.

### Social support and cumulative adversity

In terms of social support for workplace violence, participants reported a lack of clarity on what meets the criteria for violence, especially for sexual harassment from male patients towards female staff.

> “It wasn’t until right at the end that another senior member of staff experienced the sleazy looks, the looking up and down, staring at the chest but no one recognised that as the behaviour of falling under violence, aggressive, harassing abusive behaviour. It’s like people only interpret it as you being physically touched by a patient […] or someone shouting in your face, that seems to be the benchmark.” Participant 3, Hospital Nurse, White British.

Following violent incidents in the workplace, there was a perceived lack of support from senior management, which exacerbated beliefs that nothing would be done and, paradoxically, left some staff feeling like they had done something wrong by reporting it. Some participants described being actively discouraged from reporting the incident and/or told that the instance of workplace violence was their own fault.

> “When I raised the sexual harassment they [managers] first ignored it and second when I didn’t let them ignore it they tried to blame me in that they said ‘well you had a very quiet day that day so, you know, you should have been able to prepare and manage it better to prevent it from happening’.” Participant 9, Hospital Nurse, White British.

Participants were affected by the cumulative experience of violence both in and outside the workplace. For example, sexual harassment during the commute to work lowered one’s resilience to deal with other forms of violence in the workplace.

> “I just get on my bike and then cycle here but still then most mornings like today you get van drivers slowing down, driving next to you, staring at you and that happened this morning. It happens most days … You try not to lose focus and you just carry on but […] it must lower resilience before you’ve even got to work but I don’t think we acknowledge it.” Participant 3, Hospital Nurse, White British.

### Mental and physical health outcomes

The psychological consequences of workplace violence included feeling anxious, stressed and burnt out. Hypervigilance and anticipation of further violence were also commonly reported, and created an internal conflict between wanting to avoid a particular patient and one’s duty of care which is upheld by the Nursing and Midwifery Council (NMC) Code of Conduct.

> “With many patients, when they come into A&E and they get referred to you, and automatically you just get that knot in your stomach that you are going to have to go and assess somebody who you know is going to be quite difficult or quite threatening.” Participant 5, Hospital Nurse, White and Black Caribbean.

> “I mean, in a way you kind of want to avoid going there, you know, which sounds horrible, because as nurses we should provide care regardless of, you know, everything, but I was nervous and I didn’t want to be allocated for that visit.” Participant 11, Community Nurse, Mixed Ethnic Background.

Participants also felt scared, angry, undervalued and traumatised, which had far-reaching effects on both their professional and personal lives, and ultimately led to them wanting to leave the nursing profession.

> “I don’t want to work for them anymore. I don’t want to be here anymore. I am … [Gets upset] Like really traumatised by it and I don’t know what to do. And I feel like it follows me in every other aspect that I have in my life and I feel like it makes, it just made me constantly sad and flat and anxious. And I feel like I am a constant boring downer on all of my situations, on all of my relationships and I really hate that because at the end of the day I’m actually trying to help people.” Participant 9, Hospital Nurse, White British.

As well as the impact of individual violent incidents, participants also discussed the cumulative impact that experiencing multiple violent incidents over time has on their mental health and resilience.

> “I think you might get used to it but I think the impact doesn’t necessarily get less. I think actually your resilience keeps lowering and lowering […] Then, just as your resilience improves, it happens again […]. It can stay on your mind a lot, it’s affected sleep, it’s affected my going into work.” Participant 3, Hospital Nurse, White British.

Finally, the physical consequences of being assaulted by patients included exacerbating pre-existing injuries, broken bones, neck injuries and being put at risk of infections such as HIV and hepatitis. Some injuries were so severe that they prevented the individual from working.

> “I have had my wrist broken, I have had a neck injury and more recently I was injured in a restraint, and I was off work for five months on crutches unable to walk as a result of that.” Participant 6, Hospital Nurse, White British.

## Discussion

In this paper, we have described a conceptual stress process model of violence exposure for nurses. The model was informed by data from semi-structured qualitative interviews with 12 hospital-based and community nurses in London. First, we described the social context in which female nurses from different ethnic groups are exposed to community violence. Second, we identified the types of workplace violence experienced or witnessed by hospital-based and community nurses. Third, we explored nurses’ perceptions of the factors contributing to workplace violence. Finally, we examined how cumulative violence impacts mental and physical health outcomes.

As outlined in our conceptual model, primary stressors, such as workplace violence, can lead to secondary stressors, such as chronic role strain, through a process of stress proliferation [33]. Participants who had experienced workplace violence reported decreased job satisfaction which caused them to leave a particular ward/service, concerns about working and living in the same area after being threatened by a patient and the adoption of personal safety measures such as avoiding public transport. Based on previous research on chronic role strain, we propose that workplace violence impacts mental and physical health directly and indirectly by adversely altering the more enduring aspects of one’s job [33, 41, 42]. We are socialised to attach importance and invest heavily in our jobs, especially if they contribute to the healthy functioning of society, as healthcare services do [43]. Having to deal with workplace violence daily, as part of one’s job, creates high levels of stress within the institutional roles that are of great importance to both the individual and society.

Furthermore, a unique aspect of this study was that the interviews were conducted in August 2021, approximately 18 months after the COVID-19 pandemic began. Participants reported that the increased burden on the healthcare system caused by the pandemic left patients more frustrated than usual which increased levels of violence to the point where existing de-escalation techniques were less effective. Previous research shows that the surgery backlog generated by COVID-19 has worsened waiting times, leading to frustration for patients and increased workload for staff [44, 45]. As a consequence, it is unclear how protocols for managing workplace violence that were developed before the pandemic and the cost-of-living crisis can be implemented now.

The extent to which a primary or secondary stressor affects an individual’s mental and physical health depends on their social and personal resources [33]. In terms of social resources, participants reported a lack of clarity on what meets the criteria for violence and a lack of support from senior management following a violent incident. In our model, we emphasize that the lack of clarity on what meets the criteria for violence not only exists within institutions but is a wider societal issue that has implications for the measurement of violence. There is evidence that rates of exposure to community violence vary depending on the definition of community violence used, with higher rates when using a broader definition of violence to include knowing victims of violence and witnessing violence [46]. Our current measures of violence both in the community and the workplace may underestimate the scale of the problem and miss out victims of violence who need support. Furthermore, the majority of studies investigating violence against nurses focus mainly on the prevalence of first-hand experiences of violence, with limited research on witnessing violence. There is research evidence, however, that witnessing violent incidents in the past is associated with holding attitudes that justify violence in the present [47]. These findings suggest that witnessing violence towards healthcare staff has social consequences that may lead to the normalization of violence towards nurses.

In terms of personal resources, participants were affected by the cumulative experience of violence both in and outside the workplace which lowered resilience to deal with violence in the workplace and exacerbated the demands of work. Previous research has shown that cumulative violence exposure is associated with increased risk of mental health problems [48]. In a sample of women attending their initial rape forensic exam, 36% had been victims of domestic violence and 60% had been victims of prior rape [49]. In a 6-month follow up interview, 6% reported another rape and 17% reported a new physical assault. These findings suggest that women and girls experience multiple types and occurrences of violence throughout their lives. When assessing violence exposure, it is important to consider the different types and sources of violence an individual experiences throughout the life course, and their history of exposure to other types of violence. A longitudinal approach to assessment and support must be taken to prevent further violent incidents and provide appropriate long-term care.

Participants were recruited from NHS Trusts across London and those who volunteered to take part in the study were female and from a range of ethnic minority backgrounds. They reported that differences in gender, age and ethnicity between patients and staff contributed to the occurrence of violence. Participants described how patients from “other cultures” did not like to be treated by female staff, especially younger nurses who were disrespected and presumed to lack the experience needed to provide appropriate care. As well as sexism and ageism from patients towards staff, there were instances of racism whereby some White patients did not want to be treated by a Black nurse. These findings suggest that gender power relations and their intersectionality with other factors, such as age and ethnicity, undermine female health workers and increase their risk of workplace violence [50].

Furthermore, the nurses in this study were living and working in urban areas of London with high crime rates [51]. In the UK, there is evidence that people from ethnic minority backgrounds are more likely to be victims of crime compared to their White counterparts [52]. Both within and outside the UK, female nurses are further subjected to the wider societal issue of violence against women and girls. The implication of these societal factors is that female nurses, particularly those from ethnic minority backgrounds, may be vulnerable to the cumulative effects of violence both in and outside the workplace. The NHS workforce is now more diverse than at any point in its history [53] and needs to provide tailored support to meet the needs of staff with multiple social identities at the intersection of gender, age and ethnicity, to not only protect against adverse health outcomes but also improve recruitment and retention of healthcare staff.

Violence prevention training is the primary intervention for addressing workplace violence and is useful in providing staff with skills in personal safety, as well as information on the ethical, professional and legal aspects of dealing with violence that happens in the workplace. However, the evaluation of these training programmes has shown weak or limited evidence of their effectiveness [54-56]. Improving the efficacy of violence prevention interventions requires a shift towards a multi-factorial approach which involves going beyond the idea of attributing violence to individual factors, such as the patient’s mental illness or frustration, and looks instead at the social and institutional context in which violent incidents occur. This approach moves the focus of interventions from the individual level (e.g., security measures to protect staff from violent individuals) to the institutional level, such as reviewing the organisational policies, practices and procedures that foster an environment in which violence is generated and perpetuated [57]. In practice, this involves a closer examination of the factors precipitating a violent incident, how it unfolds and how it is dealt with afterwards at the level of the individual staff member, the team or service they work in and the Trust as a whole.

### Strengths and limitations

There is limited qualitative research on the mental and physical impact of workplace violence for nurses from ethnic minority backgrounds in the UK, especially those working in the community. Furthermore, some studies are limited by only assessing direct experiences of violence and fail to recognise that indirect experiences such as witnessing and hearing about violent events have a substantial impact on mental health symptoms, particularly among young women and girls [58, 59]. This study is the first qualitative study to capture the experiences of violence, experienced or witnessed, among nurses from different ethnic groups in both hospital and community-based settings. However, nurse participants were recruited from two NHS Trusts in south London and only female nurses volunteered to take part so findings may not be representative of male nurses or nurses from other parts of London or beyond. Furthermore, the voluntary nature of study participation gives rise to self-selection bias. The nurses we approached were not required to provide a reason for declining participation in the study so it is possible that those who took part differed from those who did not, in terms of motivation or workplace/personal experiences of violence. While bank (temporary) staff were excluded from the study, some participants reported witnessing higher levels of aggression towards bank staff compared to permanent staff which calls for further investigation into the role that employment status plays in workplace violence for nurses. Interviews took place in August 2021, approximately 18 months after the start of the COVID-19 pandemic, providing unique insights into the impact of the pandemic on levels of workplace violence.

## Conclusions

Hospital-based and community nurses from different ethnic groups are exposed to violence both in the community and the workplace with serious negative consequences for mental and physical health, particularly for those exposed to multiple violent incidents. NHS Trusts must improve access to mental health and well-being resources to staff affected by workplace violence, particularly for those who hold multiple intersecting social identities. Furthermore, we need to move away from interventions that put the onus on nurses to protect themselves or focus only on the physical safety measures in hospitals, and move towards interventions addressing the social and institutional factors that put nurses at risk of violence exposure and/or create an acceptable climate for violence. Future research should expand their criteria for violence to include witnessing or hearing about violent incidents and take a longitudinal, qualitative approach to understanding the consequences of violence.

## Data Availability

All data produced in the present study are available upon reasonable request to the authors

## Acknowledgements

The study was conducted by King’s College London (KCL) in partnership with NHS Race and Health Observatory (www.nhsrho.org). SH leads the TIDES study funded by Wellcome [203380/Z/16/Z] and the Economic and Social Research Council (ESRC) [ES/V009931/1]. SH is supported by the ESRC Centre for Society and Mental Health at KCL [ES/S012567/1] and the NIHR Biomedical Research Centre at South London and Maudsley NHS Foundation Trust.

